# Oncofetal Protein CR-1 in a new clinical role: a potential tumor marker for Oral Squamous Cell Carcinoma

**DOI:** 10.1101/19000950

**Authors:** Jain Anu, Mallupattu Sumanth Kumar, Thakur Reetu, Mohindra Satyawati, Bal Amanjit, Das Ashim, Ghoshal Sushmita, Pal Arnab

## Abstract

**PURPOSE:** CR-1 (CR-1) is an oncofetal protein with its role as a key factor in early process of carcinoma has been evaluated in cases of various cancers. However, very few studies have reported its role in oral cancers, which is the sixth most common cancer around the world, particularly with high prevalence in developing countries. Oral squamous cell carcinoma (OSCC) is the most predominant (90%) of all the histological types of oral cancer. Late detection, associated with increased morbidity and mortality, is mainly attributed to non-availability of a suitable biomarker for the disease. In the present pilot study we have evaluated the role of soluble CR-1, in serum as a potential tumor marker for OSCC.

**METHODS:** CR-1 was estimated using sandwich ELISA in serum samples of 50 biopsy proven OSCC patients (pre and post treatment) along with age and gender matched healthy controls. Immunohistochemistry was also done in corresponding tumor tissue sections to check the expression of CR-1.

**RESULTS:** Pre-treatment CR-1 was found to be 2.25 fold higher in serum of OSCC patients as compared to control (p<0.0001***), which was reduced to 1.6 folds post treatment (p=0.0006***). CR-1 levels were comparatively higher in early stage of disease. Upon IHC 80% of the cases were found to be positive for CR-1.

**CONCLUSION:** This study provides evidence that serum levels of CR-1 are elevated in patients of Oral Squamous Cell Carcinoma, which decrease post treatment. Also, the association of expression of protein with tumor progression predicts CR-1 as a molecule that can be further evaluated as a potential tumor maker in OSCC.

## 1. Introduction

Oral Squamous Cell Carcinoma (OSCC) accounts for about 90% of neoplasm originated from oral cavity and is the sixth most prevalent cancer in the world [1]. The annual estimated incidence is around 300,000 globally of which 62% cases occur in developing countries [2]. The risk factors associated with the disease are smoking, alcohol consumption or tobacco consumption [3–5]. A significant genetic-environmental interaction and viral association such as HPV have also been marked as potential risk factors [6–13].

Despite the fact that oral cavity is easily accessible for regular clinical examinations, OSCC is mostly detected at late stages [14]. Tissue biopsy with histopathological evaluation is the most reliable method for diagnosis of OSCC till date [15, 16]. With the progress in the medical and basic sciences, there has been improvements in the management strategies with better understanding of the pathogenesis of OSCC at molecular level. However, the survival rates have not improved over the last several decades [17], mostly attributed to non-availability of suitable tumor marker.

Thus, identification of a potential tumor marker for OSCC is definitely be beneficial to diagnose the disease at earlier stages and presumed to result in better outcomes.

Cripto-1 (CR-1) is an oncofetal protein encoded by gene located on chromosome 3p21 [18]. It belongs to EGF-CFC (epidermal growth factorCR-1-FRL-1-Cryptic) family [19]. CR-1, like other EGF-CFC proteins, found to play a role in embryogenesis, where it functions along with nodal, TGF-β (transforming growth factor-β) and GDF-1/GDF-3 (Growth and differentiation factor 1 and 3) as their co-receptor [20]. During embryogenesis it is found to be involved in gastrulation, specification of endoderm and mesoderm, cardiogenesis etc. [21]. *In vitro* studies, reporting induction of transformation of a variety of cells by overexpressed CR-1, provide the evidence of its oncogenic activity [22, 23]. Structurally, CR-1 consists of four major domains which are; a signal peptide domain at amino terminal, an Epidermal Growth Factor-like domain, a cysteine-rich domain and a short domain, present at carboxyl terminal (hydrophobic in nature), containing sequence for glycophosphatidylinositol (GPI) anchorage and cleavage [20]. The GPI anchorage and cleavage sequence is responsible for existence of CR-1 in two distinct forms, i.e. membrane bound form and soluble secretory form. Although both the forms have been found active in tumorigenesis, membrane bound form, is proposed to be the key player in this process. Various studies have shown involvement of membrane bound CR-1 in process of early carcinogenesis, growth and survival of tumor tissue, metastasis etc. [21]. Studies carried out in a number of tumor tissues of various biological sites viz; breast, colon, gastric, lung, liver suggest this role of membrane bound CR-1 [24]. On the other hand, soluble/secretory form has been proposed to be related to the disease progression and prognosis.

There is only one study showing relevance of CR-1 in Oral Cancer. Yoon H.J. et al, in 2011, reported high expression of membrane bound CR-1 in tumor tissue of oral squamous cell carcinoma (OSCC) patients [25]. However, the study did not include the expression analysis of soluble form of CR-1. As the estimation of soluble CR-1 from serum is much easier by using methods like ELISA than IHC from tumor tissue as reported by Yoon HJ, it may be used for repeated measurements aiming to the response to the therapy as a prognostic marker.

In present pilot study we evaluated soluble form of CR-1 from serum of the histologically diagnosed patients of OSCC as a potential diagnostic and/or prognostic marker for the first time.

## 2. Materials and methods

### 2.1 Patients and specimen collection

After ethical approval by Institutional Ethics Committee, patients and healthy volunteers were included in the study after obtaining written informed consent. Fifty biopsy proven OSCC patients attending the Department of Radiotherapy were included in the study. Patients intended for curative radical radiotherapy with 60Gy fractionated doses were included in the study. As there is no prior scientific literature available regarding the levels of soluble CR-1 in serum of OSCC patients, we planned the study to be pilot study and included 50 consecutive patients matching our inclusion and exclusion criteria. Patients received prior treatment or planned for palliative treatment were excluded. Blood samples from patients and healthy controls (age and sex matched) were collected. Serum was separated and stored in - 80°C for further use for estimation of soluble CR-1 by ELISA. Histological biopsy sample of the corresponding patients were also processed for confirmation of diagnosis as well as immunohistochemistry (IHC) to check expression of membrane bound CR-1 levels.

### 2.2 Clinical parameters of the patients

Clinical parameters of each patient were noted including age, gender, tumor differentiation, T stage, N stage and TNM stage. All the patients were followed up for their grade and stage of disease and disease relapse (if any) and survival were recorded on the basis of follow-up.

### 2.3 ELISA

A sandwich type serum ELISA was done using DY145 ELISA kit for CR-1 by R&D system™ USA, following manufacturer protocol. Briefly, 4μg/mL of capture antibody was coated in a 96-well plate. 100μL of neat serum samples followed by 500ng/mL of detection antibody were incubated at room temperature for 2 hours respectively. Signal detection was performed by incubating streptavidin-HRP and colorized by ortho-phenylenediamine substrate solution (Hi-media, MB-204, India). Human recombinant CR-1 (provided in the kit) was used as standard. The absorbance was measured at 450 nm to determine the colorimetric signal.

### 2.4 Immunohistochemistry

Parafinized tissue section was fixed on poly-L-Lysine coated glass slides. After deparafinization, endogenous peroxidase activity was blocked. Further, antigen retrieval was undertaken by boiling in citrate buffer (pH 6.0) for half an hour. The CR-1 antibody (ab19917 Abcam, USA) was used. Detection was done by HRP conjugated antibody by DAKO (Real Envision™ Denmark) and colorized by DAB. Pancreatic tissue was used as positive control. The results IHC were analyzed by two independent observers.

### 2.5 Statistical analysis

For statistical analysis, GraphPad Prism 6 software was used. Mann Whitney test and Wilcoxon matched pair test were used to check the significance of the results, p ≤ 0.05 is taken to be significant for all tests.

ROC curve analysis was done to find out the specificity and sensitivity of the test.

## 3. Results

Fifty histologically confirmed OSCC patients were included in the study. Most of the patients were male (49, 98%) with age between 30-80 years (median ∼54 years). The control population involved was age and gender matched **(Table 1)**. Clinical stage was T_1_/T_2_ in 46% (n=23) of cases and T_3_/T_4_ in 54% (n=27) cases; N_0_/N_1_ in 60% (n=30) cases and N_2_/N_3_ in 40% (n=20), however none of the cases enrolled in our study had any distant metastasis.

**Table 1:** Characteristics of cases and controls involved in the study

|  | Cases | Control |
| --- | --- | --- |
| Number (n) | 50 | 50 |
| Age Group (years) | 30-80 (median 54) | 24-60 (median 48) |
| Gender | 98% male | 90% male |
| Mean CR-1 levels (pg/mL) | Pre-treatment- 365.10 | 167.36 |
|  | Post-treatment -260.18 |  |

### 3.1 High levels of serum CR-1 in OSCC patients

We have previously shown that serum CR-1 levels of cancer patients were found to be increased by 2.25-fold as compared to control **(Fig 1A)** with a mean value of 365.10pg/mL and 167.36pg/mL respectively ***(p=0.0001***)*** [26].

**Fig 1:**
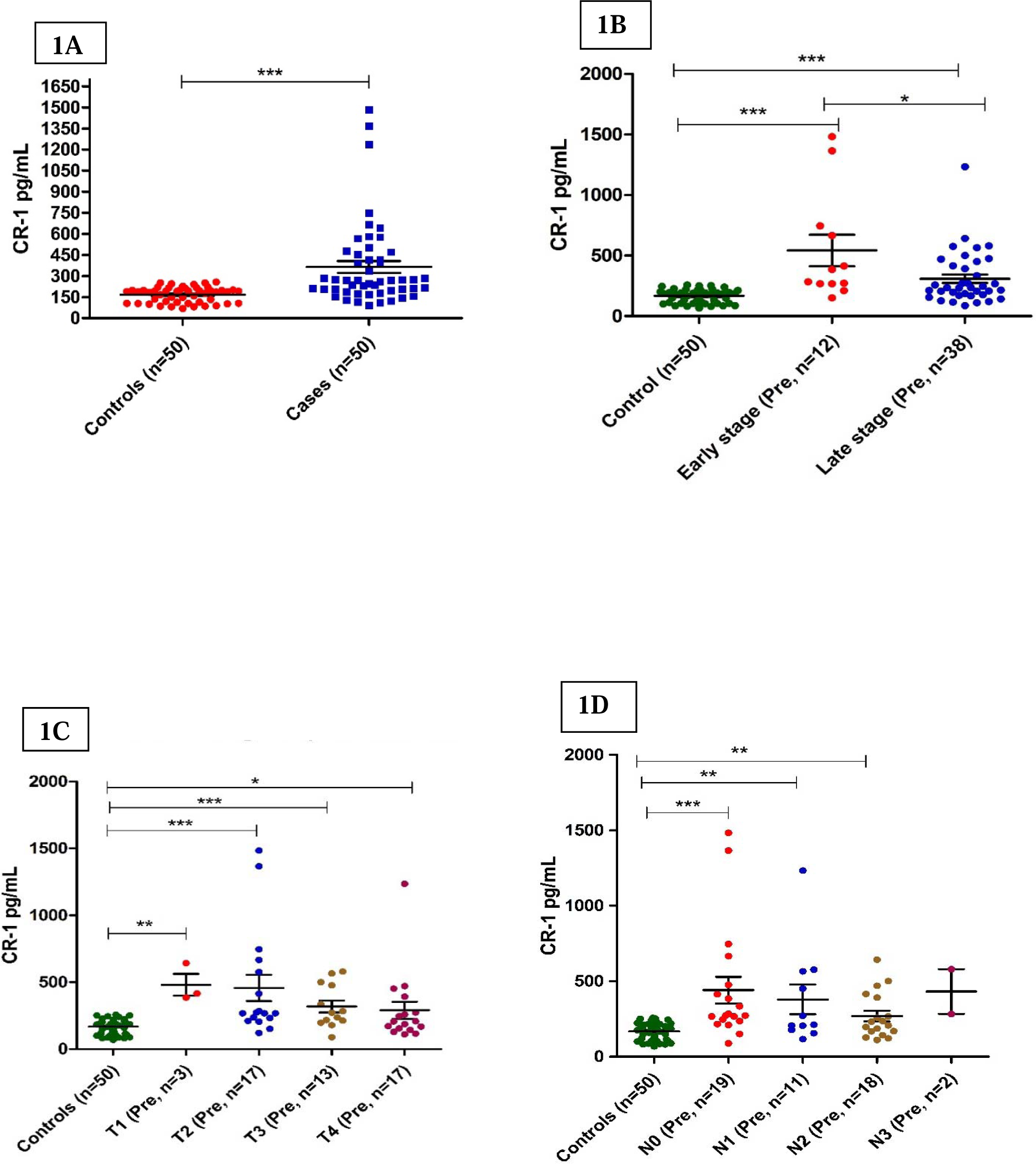
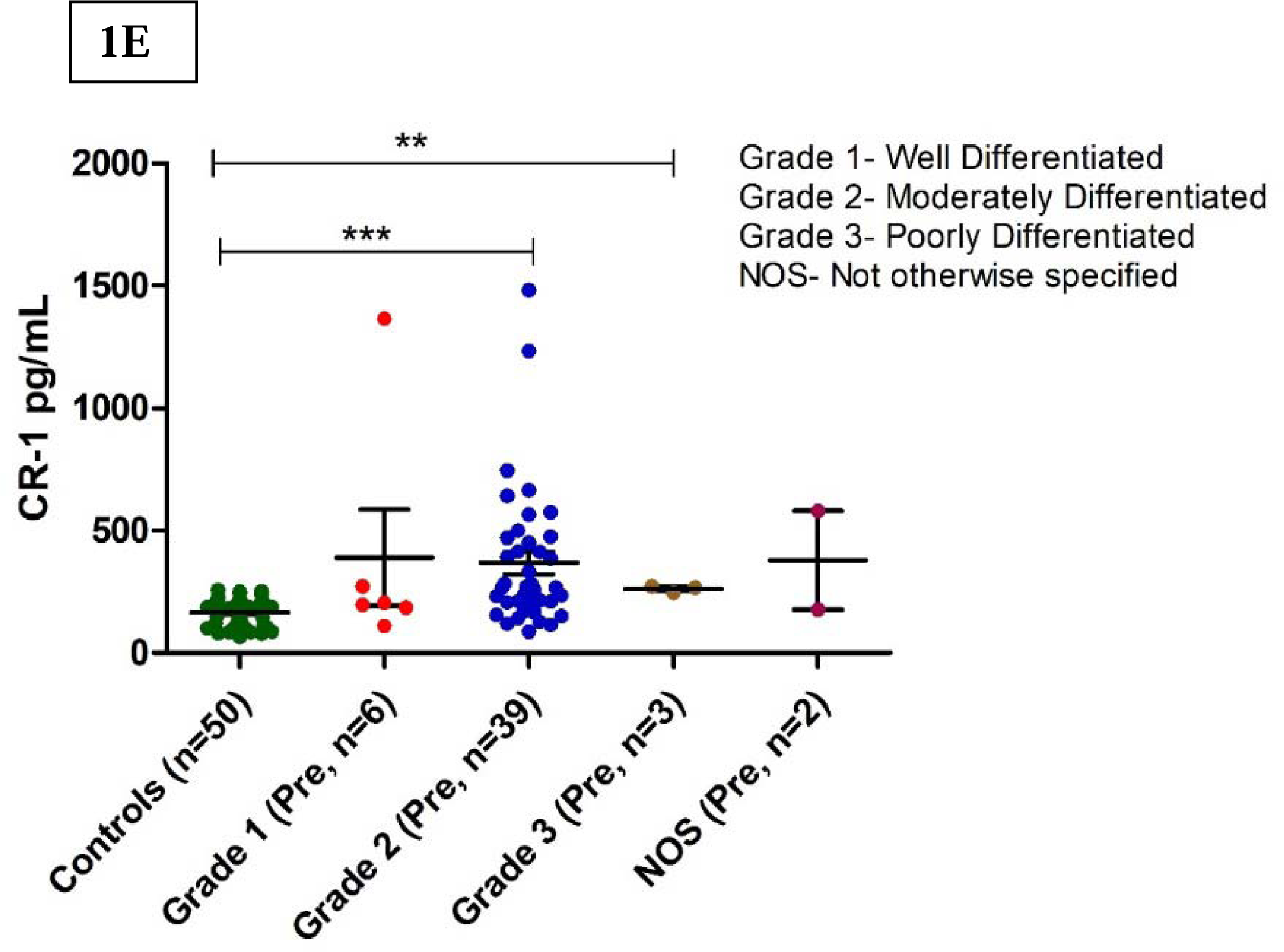
**(A)** Increased level of CR-1 expression were observed in cases as compared to control. Expression was also increased **(B)** in early and late stages of cancer with respect to control, **(C)** in all the stages as per tumor size, **(D)** as per lymph nodes involvement and **(E)** in different histological grades.

#### CR-1 levels and association with clinical variables

Association between serum CR-1 levels and cancer staging and grading was also evaluated. CR-1 levels were found to be increased significantly both in early (T_1_&T_2_; ***p<0.0001\*\*\****) and late stage (TNM classification) of the disease (T_3_&T_4_; ***p<0.0001\*\*\****). Moreover it was also found that there was more increase in CR-1 level in early stage (T_1_/T_2_) than the late stage (T_3_/T_4_) **(Fig 1B)**. CR-1 levels were increased significantly in all the stages as per tumor size (T stage) i.e., T_1_ (***p=0.0021\*\****), T_2_ (***p<0.0001\*\*\****), T_3_ (***p=0.0002\*\*\****) and T_4_ (***p=0.0204\****), nodal involvement (N stage) i.e. N_0_ (***p<0.0001\*\*\****), N_1_ (***p=0.0029\*\****) and N_2_ (***p=0.0045\*\****) and histological grading i.e. moderately differentiated (***p<0.0001\*\*\****) and poorly differentiated (***p=0.003\*\****)(**Fig. 1C, 1D & 1E**).

### CR-1 levels decreased in patients post treatment

The mean level of CR-1 in patients which was 365.10pg/mL decreased to 260.18pg/mL after treatment with radiotherapy and/or surgery suitable for grade and stage of tumor **(Fig 2A)** with a significance level of ***p=0.0002\*\*\****.

**Fig 2:**
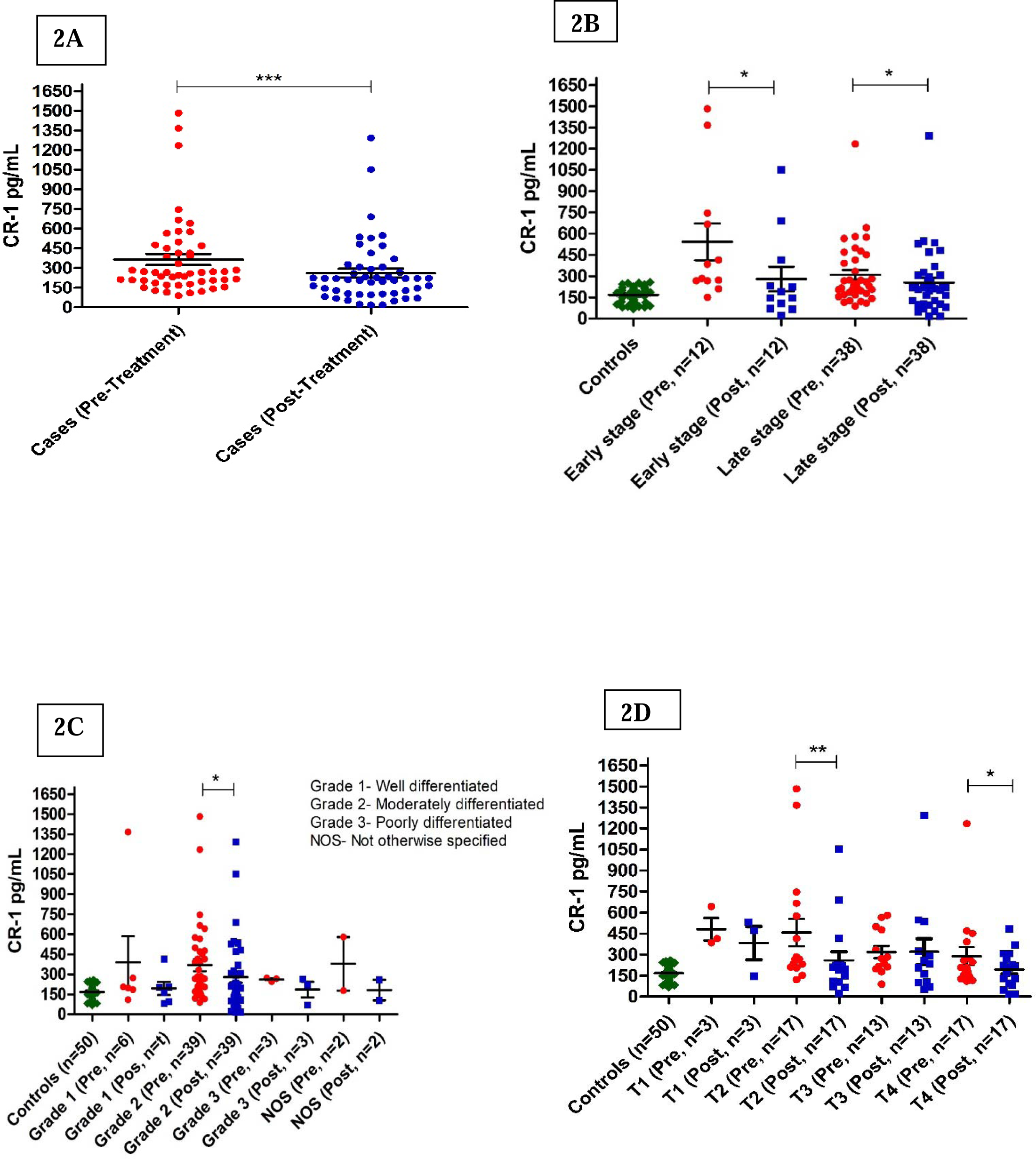
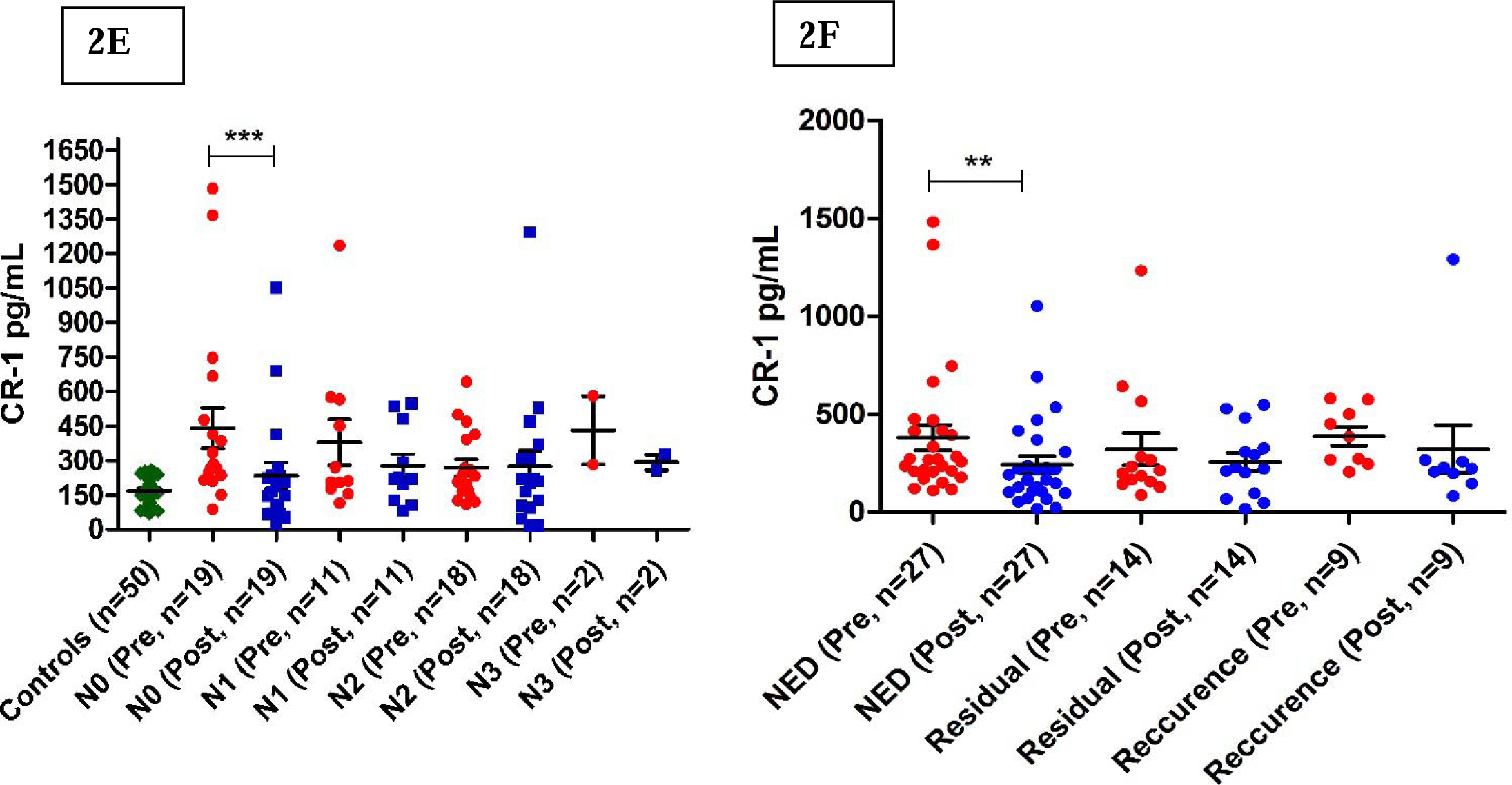
**(A)** CR-1 levels decreased significantly upon treatment. **(B)** CR-1 levels decrease post-treatment in both early and late stages of the disease. **(C)** Post treatment levels of CR-1 in different histological grading. Decrease in levels of CR-1 was observed upon treatment **(D)** in all the stages as per tumor size and **(E)** as per lymph nodes involvement. **(F)** CR-1 levels in cases as per the disease status upon completion of treatment.

Both, early (T_1_&T_2_; ***p<0.01\****) and late stage (TNM classification) of the disease (T_3_&T_4_; ***p<0.01\****) showed significant decrease in CR-1 levels upon treatment **(Fig 2B)**. Though the levels decreased in all the histological grades, however only in moderately differentiated histological grades of tumor tissue, levels were significantly decreased post treatment ***(p=0.03*)* (Fig 2C)**.

Increased CR-1 levels were associated with all the stages of tumor size (T staging) and lymph nodes involvement (N staging). The decrease in levels were found in all the stages of tumor size (T staging) and lymph nodes involvement (N staging) post treatment, however the decrease was statistically significant only in earlier stages i.e. T_2_ stage (***p=0.0024\*\****) and N_o_ stage (***p=0.0010\*\*\****) **(Fig 2D & 2E)**.

### CR-1 levels do not decrease in patients with post treatment residual disease

Upon follow-up patients were divided into two categories based on their disease status, one group included patients with no evidence of disease (denoted as NED) after treatment by clinical and radiological examination and the other included patients with residual disease (denoted as RD) after treatment. It was observed that patient group with no evidence of disease reflected significant decrease in CR-1 levels after treatment with respect to their pre-treatment status. However, decrease in post treatment CR-1 values in residual disease group was non-significant **(Fig 2F) (Table 2)**.

**Table 2:** Pre and post treatment CR-1 levels in cases with different classifications as per disease staging, grading and post treatment disease status.

| Characteristic | Before treatment<br>CR-1 levels (pg/mL) | After treatment CR-1<br>levels (pg/mL) | p value |
| --- | --- | --- | --- |
| **TNM Classification** |  |  |  |
| Early stage (T<sub>1</sub> & T<sub>2</sub>) (n=12) | 542.27 | 280.3 | **0.011\*** |
| Late Stage (T<sub>3</sub> & T<sub>4</sub>) (n=38) | 309 | 253.8 | **0.014\*** |
| **T Stage (Tumor Size)** |  |  |  |
| T<sub>1</sub> (n=3) | 480.7 | 381.7 | 0.25 |
| T<sub>2</sub> (n=17) | 456.7 | 258.5 | **0.0024\*\*** |
| T<sub>3</sub> (n=13) | 318.2 | 321.5 | 0.2487 |
| T<sub>4</sub> (n=17) | 288.9 | 193.5 | **0.0443\*** |
| **N Stage (Lymph Node Involvement)** |  |  |  |
| N<sub>0</sub> (n=19) | 441.1 | 234.3 | **0.001\*\*\*** |
| N<sub>1</sub> (n=11) | 379.1 | 276.5 | 0.1826 |
| N<sub>2</sub> (n=18) | 268.9 | 273.9 | 0.1979 |
| N<sub>3</sub> (n=2) | 431.5 | 292.4 | 0.2929 |
| **Histological Grading** |  |  |  |
| Well Differentiated (n=6) | 389.7 | 194.2 | 0.0781 |
| Moderately Differentiated<br>(n=39) | 368.5 | 280.1 | **0.034\*** |
| Poorly Differentiated (n=3) | 280.1 | 262 | 0.1833 |
| **Disease Status Post Treatment** |  |  |  |
| No Evidence of Disease<br>(n=27) | 380.4 | 241.9 | **0.0056\*\*** |
| Residual Disease (n=14) | 321.4 | 256 | 0.1629 |
| Recurrence (n=9) | 387.12 | 321.53 | 0.08 |

### CR-1 was overexpressed in the tissue of OSCC patients

In addition to estimation of soluble CR-1 in serum, we also checked the tissue expression of CR-1 by immunohistochemistry for expression of membrane bound non-soluble component CR-1. **(Fig 3)**. 80% of the cases were positive for the CR-1 expression at tissue level. 50% of the cases were weakly positive for CR-1 expression, 23% and 7% cases showed moderate and strong expression respectively. 19% cases were found to be negative for CR-1 expression.

**Fig 3:**
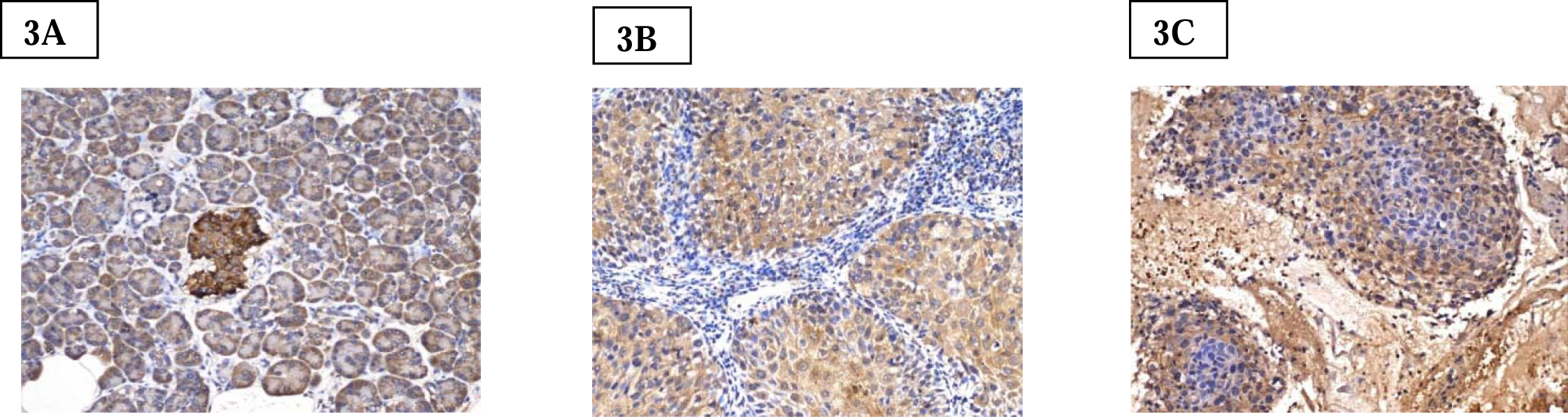
Membrane boundCR-1 expression in tumor tissue upon Immunohistochemistry. **(A)** +3 positive control (pancreas). **(B)** OSCC tissue (+2, 80% tumor). **(C)** OSCC tissue (+2, 10% tumor).

### ROC curve analysis

ROC curve analysis showed that CR-1 shows a sensitivity and specificity of 74% and 78% respectively at a cut off of 202.5pg/mL. Area under the curve was 0.80 with a statistical significant value of **p<0.0001\*\*\*** [26].

### Survival curve analysis

The patients were followed-up from the time of their enrollment in the study till the completion of the study. The overall survival and disease-free survival of the patients with CR-1 values below and above 202.5 pg/mL (cut off value) was compared. However, the data was not significant statistically (data not shown).

## 4. Discussion

Although CR-1 plays a key role during embryogenesis, it ceases to express in all tissues barring some, like developing mammary glands [27], soon after birth. However, differential expression of CR-1 in various cancerous cells like colon carcinoma, breast cancer, gastric cancer, glioblastoma, etc. has also been reported. Despite the evident literature contributing to the understanding of mechanism and expression of CR-1 in various tumor tissues, expression status of this oncofetal protein has not been explored much in OSCC, except by Yoon H.J. et al in which author studied only the membrane bound expression of CR-1 in OSCC tumor tissue [25]. While membrane bound CR-1 has already been shown to be associated with OSCC, it was hypothesized that the soluble component, which is released in serum after cleavage of GPI linkage, can be more easily estimated in serum than tissue. Moreover, repeated estimation before and after the treatment as well as during follow up makes soluble CR-1 a better suitable candidate as a marker than its membrane bound counterpart.

We found that serum CR-1 levels were increased by more than two folds in OSCC patients as compared to healthy controls with a p value of **<0.0001\*\*\***. These results are consistent with the previous finding for other cancers like breast and colon cancer [28]. Significant decrease in the CR-1 levels **(p<0.0006***)** upon successful treatment supports the fact that OSCC tumor population is significantly rich for the expression of soluble CR-1. This is also supported by the fact that there was minimal or no decrease in cases of remaining residual diseases after therapy. The exact role of CR-1 in oral oncogenesis cannot be concluded from our study, however it is suggested that it might be playing an important role as evident by the increased abundance of both membrane bound and soluble component of CR-1. *In vitro* and *in vivo* studies have already been conducted in the past to show the oncogenic role of CR-1 in cell proliferation, migration, invasion and tumor angiogenesis during tumorigenesis [29, 30]. Epithelial to mesenchymal transition, a crucial step for metastasis of tumor cell has also been shown to involve CR-1 as a key player. The same is postulated to be true in cases of OSCC also.

Correlations of serum CR-1 levels with clinical variables reveal that higher CR-1 levels are associated with earlier disease conditions. It was observed that serum CR-1 levels were more increased with respect to the control group in early stage (TNM classification) cohort of patients (fig 1B). Similar pattern was observed for staging based on tumor size (T staging) and lymph nodes involvement (N staging) (fig 1C & 1D). This suggests that soluble CR-1 is more active and functional during early stages of carcinogenesis. It has been reported in other cancers that CR-1 is being expressed in cancer stem cell population, termed as cancer stem cell marker, and this population of cancer cell is involved in progression and invasion of cancer tissue, in the early stage of cancer [19]. Our observation of association of higher serum CR-1 levels in early disease conditions can be explained by this.

Immunohistochemistry of the OSCC tumor tissue, in our study, reveals that 80% of the cases showed positivity for CR-1. This expression is more than that reported earlier for OSCC in literature [25]. However, we could not find significant correlation between tissue expression of CR-1 with tumor size, lymph node metastasis, clinical stage, or recurrence as also reported by Yoon H.J. et al [25].

OSCC being the sixth most common cancer worldwide and third most common in developing countries like India is an important health issue and the poor 5-year survival rate associated with the disease, exacerbate the scenario. Multiple risk factors like tobacco consumption (smoking or chewing), betel quid consumption, and alcohol consumption are highly prevalent in the developing countries, which increased the risk of OSCC. Oral cavity, in-spite of being easily available to access and review for disease symptoms, the disease is not detected in early stages in most of the cases. This is because the symptoms are often too vague to be considered as cancer or in other cases ignored due to unawareness or sometimes the phenotypical symptoms don’t appear to be noticed in physical examination. Thus, in this scenario a biomarker, levels of which can be measured easily from an easily accessible body fluid like blood, urine or saliva may be a good diagnostic and/prognostic marker of disease conditions.

Our study for evaluation of serum CR-1 levels is an effort towards identification of a potential biomarker for OSCC. Increased serum CR-1 levels which go down upon treatment and correlation with early disease conditions suggests that this can be a potential prognostic or diagnostic marker for OSCC. The survival analysis was done which was found to be insignificant and this may be explained by the small size of study population. As our study is a pilot study and proof of concept in nature with limited number of patients, further studies in larger patient cohorts with different stages are required to strengthen our observation regarding the expression pattern of the soluble CR-1 protein in OSCC. However, the study has been conducted and reported in accordance with the REMARK (REporting recommendations for tumor MARKer) criteria [31].

Moreover, the role of CR-1 in oncogenesis of OSCC also need to be explored for future prospective. This will not only help to establish a biomarker for the disease but can also help to understand the role of CR-1 in the disease progression and can help to find a therapeutic measure by targeting the CR-1 protein.

## Data Availability

All raw data is available from corresponding author on request.

## Acknowledgement

This work was supported by Department of Biotechnology (DBT), New Delhi, India under the RGY1 scheme (BT/PR6139/GBD/27/366/2012) and intramural grant (Post Graduate Institute of Medical Education and Research-71/4-Edu/11/1537). We are also thankful to Indian Council of Medical Research (ICMR) for providing fellowship to RT [3/1/3JRF-HRD-131 (10709)] and AJ [3/1/3JRF-HRD-22 (10519)].

## Notes

### Competing Interest Statement

The authors have declared no competing interest.

### Funding Statement

Department of Biotechnology (BT/PR6139/GBD/27/366/2012):Dr Arnab Pal
Post Graduate Institute of Medical Education and Research (71/4-Edu/11/1537):Dr Arnab Pal
Indian Council of Medical Research (3/1/3JRF-HRD-22 (10519)):Ms. Anu Jain
Indian Council of Medical Research (3/1/3JRF-HRD-131 (10709)):Ms. Reetu Thakur

### Author Declarations

All relevant ethical guidelines have been followed and any necessary IRB and/or ethics committee approvals have been obtained.

Any clinical trials involved have been registered with an ICMJE-approved registry such as ClinicalTrials.gov and the trial ID is included in the manuscript.

